# Global trends in occupational disease reporting: a systematic review

**DOI:** 10.1101/2024.09.19.24314032

**Authors:** Levina Chandra Khoe, Siti Rizny Fitriana Saldi, Marsen Isbayuputra, Muchtaruddin Mansyur, Virginia Wiseman, Augustine Asante

## Abstract

**Background:** Disease reporting is often unreliable and faces many challenges, making it difficult to estimate the true burden of occupational diseases, defined as any disease that is caused by the work activity or environment. This study aimed to assess the global reporting and underreporting rate of occupational diseases, and to identify the factors affecting the underreporting of occupational diseases.

**Methods:** Following the Preferred Reporting Items for Systematic Reviews and Meta-Analysis (PRISMA) guidelines, this study searched Medline (PubMed), CINAHL, EMBASE, Scopus, Web of Science, WHO Institutional Repository for Information Sharing (IRIS) database, Dimensions, and Google Scholar. We used search terms related to reporting and underreporting of occupational diseases or illnesses. The selected records were screened, and data extracted using the Covidence software tool. Screening and quality assessment were conducted by two independent researchers and finalized by a third researcher. The quality of the evidence was assessed with the Mixed Methods Appraisal Tool. This study is registered on PROSPERO, number CRD42023417814.

**Results:** A total of 121 studies from 29 countries were identified, all coming from high-income and upper-middle-income countries. The incidence rate of occupational disease varied widely, ranging between 1.71 to 1,387 per 100,000 employees yearly. The highest number of annual cases was reported in the agricultural sector (ranging from 33 to 6,431), followed by the health sector (146 to 5,508), and then the construction sector (264). Two studies evaluated rates of underreporting, which varied from 50% to 95%. The main factor contributing to underreporting was employee concerns about job security.

**Conclusions:** The results reveal a significant gap in the reporting of occupational diseases among high-income and low-middle-income countries. This review also identifies variations in reporting mechanisms across different countries. Our findings highlight the need to establish a national system for reporting occupational diseases that engages employers, employees, and healthcare providers.

## Introduction

In 2016, the World Health Organization (WHO) and the International Labour Organization (ILO) produced the first joint estimates of the work-related burden of disease and injury. They reported a total of 1.9 million deaths and a loss of 89.7 million disability-adjusted life years (DALYs) attributable to 19 occupational risk factors (1). To prevent occupational diseases, obtaining correct information on their prevalence is necessary. According to a global survey conducted by the WHO, about 93.5% of countries globally collect data on occupational diseases and 68.5% of these maintain a national registry of occupational diseases and accidents (2). The same survey also shows that about 87.2% of high-income countries (HICs) have national registries, and around 40.8% of low- and middle-income countries (LMICs) do not have these registries. Even where national registries exist, the level of registration is reported to be low, especially in LMICs (2). Concerns about incomplete registry data especially around occupational risk factors has also been raised (3).

There are recognized challenges associated with the collection of occupational diseases data in terms of population coverage, source of information, and coordination between multiple authorities (3). Firstly, the population covered by reporting systems mainly includes employees working in the formal sector, excluding the self-employed, part-time workers, casual workers, seasonal workers, and those in micro to small enterprises (4). These gaps in population coverage result in underreporting, especially in LMICs, where more than two-thirds of the population works in the informal sector (5). Secondly, to diagnose occupational disease, physicians require robust evidence on the possible occupational origins and their level of exposure (6, 7), which must be sourced from employers (8). Insufficient training in occupational medicine for physicians has also been identified as one of the factors contributing to the under-diagnosis of occupational diseases (9–11). Apart from human factors, some occupational diseases require a long latency period before the appearance of first symptoms. For instance, the latency period from exposure to carcinogenic substances for bladder cancer is about 14 years, which makes it difficult to establish the link between exposure and disease (12). Lastly, the responsibility to collect data on occupational diseases may be divided among different organizations. In many countries, the Ministry of Health and the Ministry of Labour often require this information for disease surveillance purposes and for developing occupational health programmes. Employers and insurance agencies need the information for workers’ compensation schemes(13). Each organization may use different reporting mechanisms, which could increase the complexity and fragmentation of documenting occupational diseases(2).

Occupational disease reporting systems are important sources of information for understanding disease patterns among workers in a country and for developing effective prevention programmes. However, statistics on occupational diseases are often unreliable and remain scarce in many countries, particularly in LMICs, where many workers have a higher risk of developing occupational diseases (14). While all cases of occupational disease must be reported by employers, employees, and/or physicians, many employees are unaware that their disease may be caused or worsened by the work environment (15, 16). Some may be aware but choose not to report for fear of potential repercussions, including losing their jobs (17). Factors that contribute to underreporting of occupational diseases might also originate from employers who may not consider the employee’s case as work-related. Additionally, there is a lack of enforcement of occupational health and safety regulations in many countries. A lack of strictly enforced sanctions on employers who fail to report confirmed or suspected occupational-related diseases to authorities has been documented in many countries (17–20).

Occupational diseases are often described using the “icebergs phenomenon”(21). This means that most cases may be invisible or mistakenly diagnosed as non-occupational diseases. Thus, the number of cases reported in many countries may be just the tip of the iceberg. While studies have documented the burden of occupational diseases, there has been limited effort to explore the global disease pattern and identify its reporting mechanisms. To our knowledge, no systematic review exists on the global reporting of occupational diseases. This systematic review aims to fill that gap by bringing together evidence on the reporting of occupational diseases in both high-income countries and LMICs. The review covers all types of occupational diseases. Occupational injuries or accidents are not included because they are more easily recognized and better reported compared with most diseases. Our goal is to systematically review the reporting and underreporting pattern of occupational diseases based on countries’ income status, industrial development, and types of occupational diseases. We also aim to identify the factors affecting the underreporting of occupational diseases.

## Methods

The protocol for this review was prepared according to the Center for Reviews and Dissemination (CRD) guidelines and registered on PROSPERO, under registration number CRD42023417814. The selection of studies followed the Preferred Reporting Items for Systematic Review and Meta-Analysis-Protocols (PRISMA) 2020 guidelines. The quality of the studies were assessed using the Mixed Methods Appraisal Tool (MMAT) (22). This study is registered on PROSPERO, number CRD42023417814.

### Eligibility criteria

Studies were eligible for inclusion if they examined the reporting or underreporting of occupational diseases at the country level. Eligible studies included cohort studies, cross-sectional studies or reports from national registries, national surveillance systems, workers’ compensation schemes, or national voluntary reporting schemes. As this review focused on the reporting system at the national level, studies at the district or province level were excluded. All types of occupational diseases were included; however, occupational injuries or accidents were excluded. Review articles, editorials, guidelines, case reports, and case series were also excluded, as our focus was on empirical studies. Qualitative and mixed-methods studies were included to identify the factors underlying problems of underreporting. There were no limitations on the date of publications and language. For non-English language articles, Google Translate was used to screen titles and abstracts. These articles were kept in a separate folder but not included in the analysis, to avoid language bias when appraising the articles.

### Outcome

Outcome measures include the number of reported occupational disease cases, the rate of reporting occupational diseases, the rate of underreporting or misreporting, the number of occupational disease claims, the number of cases reported by physicians, employees or employers. The underreporting rate is defined as the ratio between the number of non-reported cases and the total number of cases (reported and not reported). The rate of misreporting refers to the ratio between the number of falsely reported cases and the total number of cases. Reported cases were classified according to the countries’ income status, industrial development, and types of occupational diseases.

### Data sources and search strategy

Searches were conducted for the following electronic databases: Medline (PubMed), CINAHL, EMBASE, Scopus, and Web of Science. Searches for eligible grey literature were carried out in the WHO Institutional Repository for Information Sharing (IRIS) database, Dimensions, and Google. Additionally, the reference lists of relevant articles were screened for titles and abstracts that include key terms.

We explored different possible terms related to reporting and underreporting of occupational diseases/illnesses including “report*”, “underreport*”, “misreport*”, “surveillance”, and “capture-recapture”, and combined them with terms for occupational diseases (“occupational disease*”, “occupational illness*”, “work-related disease”, “work-related illness”). The search strategy included a combination of Medical Subject Heading (MESH) terms and free text terms. These terms were combined with ‘OR’ and ‘AND’ Boolean operators. The full search strategy is provided in S1 Appendix.

### Study selection

Studies obtained from different data sources were combined and duplicate records were removed using the Covidence systematic review software package. All records identified in the search were initially screened based on titles and abstracts. Then, we assessed the full text of selected studies according to the eligibility criteria. Study selection was performed by two independent investigators (LK and SR or LK and MI). Any disagreements were resolved through discussion with a third reviewer (SR or MI). The third reviewer was a person not involved in the study selection process.

### Data extraction

A standardized form was developed for data extraction based on the review questions. Extracted information included basic study details such as study design, country of origin, study setting, year or timeframe for data collection, participant employment characteristics (industrial sector, job type), and the outcome data (e.g., number of cases reported, reporting rate). Additionally, the countries where the studies were conducted were stratified by income status and region according to the World Bank classification (23). One reviewer (LK) extracted the data, while a second reviewer (SR or MI) checked the data for accuracy and completeness. Any discrepancies between reviewers were resolved through consensus, or by involving a third reviewer (SR or MI) to settle the disagreement in the data extraction process. Where essential information was unclear or missing, the authors were contacted for clarification. Completed data extraction forms were uploaded to the university’s (University of New South Wales) One Drive account, which was accessible only to the reviewers.

### Quality assessment

At least two reviewers (LK and SR or LK and MI) assessed and appraised the methodological quality of the studies independently using Mixed Methods Appraisal Tool (MMAT) (22). The types of studies assessed using MMAT include qualitative studies, quantitative randomized controlled trials, quantitative non-randomized studies, quantitative descriptive studies, and mixed-method studies. Five criteria were used to assess the overall risk of bias for each study type. Each criterion is given a rating of ‘yes’, ‘no’, or ‘can’t tell’. For every ‘yes’ answer, the study was given a score of between 20 and 100, with 20 being the lowest and 100 the highest. Any inconsistencies were resolved through discussion and included a third reviewer (SR or MI), if necessary.

### Analysis and data synthesis

The articles were categorized based on their content. Articles that included the number of reported cases or incidence rate of occupational diseases were grouped into one folder and analysed to address the primary objective, i.e., to estimate the global reporting and underreporting rate of occupational diseases. Descriptive data from each reviewed study were presented as narrative text or in tables. Due to the diversity in the characteristics of the studies, conducting a meta-analysis was not possible. Thus, a narrative synthesis was performed for this systematic review. A summary table showing the number of cases per year and incidence of occupational diseases per 100,000 employees was presented. Where the incidence was originally reported using a different denominator (e.g., per one million or one thousand employees), the number was adjusted to a denominator of per 100,000 employees. The results were also classified based on the type of occupational diseases in each region, and the type of occupational diseases in each industrial sector. In every study, we calculated the number of annual cases for each type of occupational disease by adding up the total cases and dividing by the total number of years covered by the study to obtain the average number of cases per year. The average number of cases per year in each study were summed up and then divided by the total number of studies. For studies discussing the factors contributing to the underreporting of occupational diseases, these articles were separated into a separate folder and analysed thematically.

## Results

### Characteristics of included studies

From the searched databases, we identified 15,397 records. Of these, 6,885 duplicates were removed. After title and abstract screening, the remaining 410 full-text articles were assessed for eligibility. Of these 410 articles, 20 studies were in other languages; 3 German, 2 French, 4 Spanish, 2 Danish, 3 Polish, 2 Italian, 2 Norwegian, and 2 Chinese. A total of 289 studies were excluded from the 410 because they did not meet the inclusion and exclusion criteria.

That left a total of 121 articles to be considered for final review, including 118 quantitative studies and 3 qualitative studies. The study selection process is shown in Figure 1. These 121 studies covered 29 countries spread across all six geographical regions of the world (Africa, Asia, Australia and Oceania, Europe, Northern America, and Latin America). The first study was published in 1990 and the most recent in 2023. While only 15 studies were published in the first 10 years between 1990 and 2000; the number of published articles remained stagnant each year, at around 6 studies annually.

**Fig 1.**
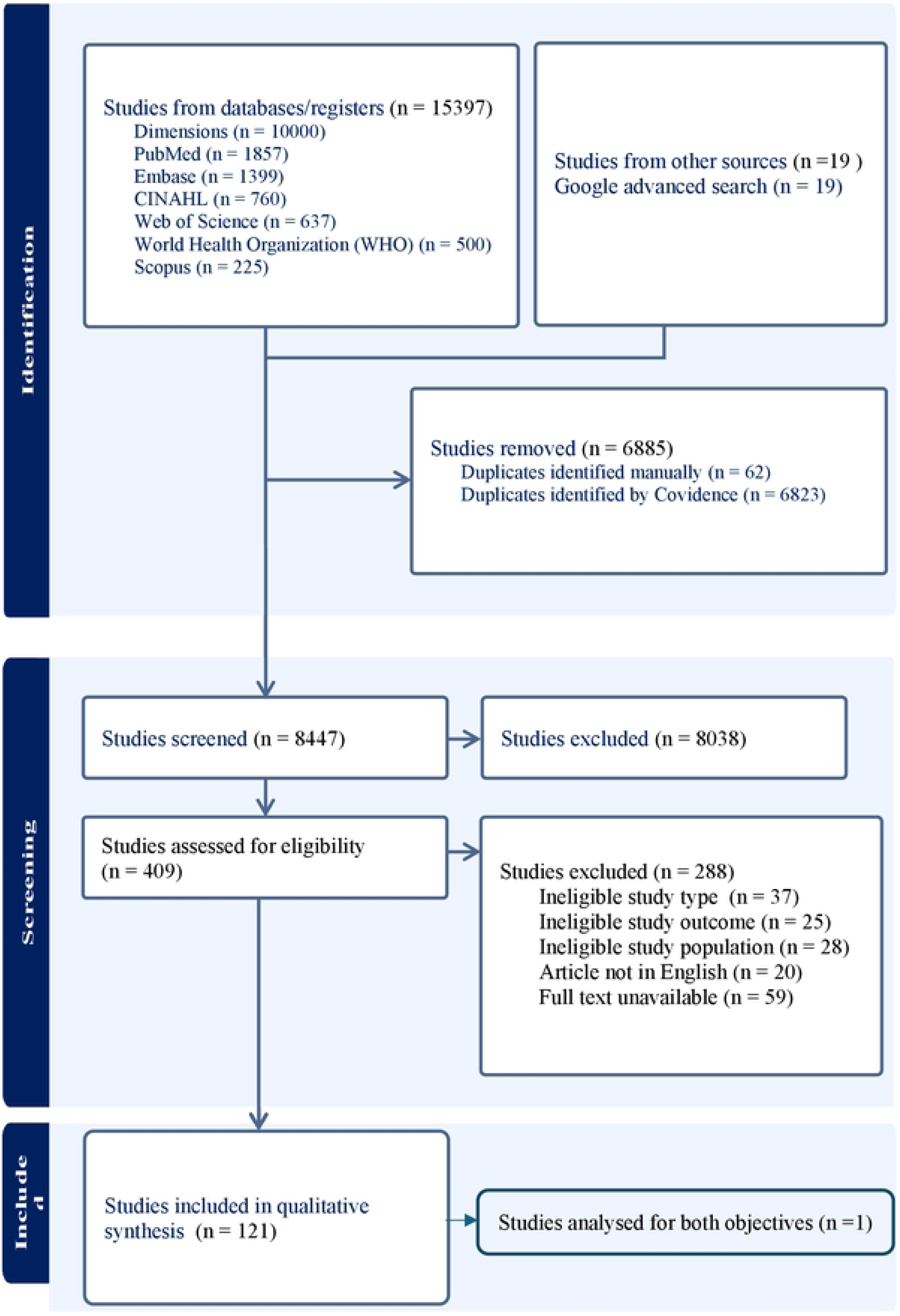
PRISMA flow diagram of study selection.

The country with the highest number of publications on occupational diseases was the United Kingdom (UK), which published 27 articles. Other top-producing countries included South Korea and Finland, contributing 9 and 8 articles respectively. Denmark, Australia, and Canada were the first countries to publish occupational disease studies between 1990 and 1996. All the articles included in this review were published in high-income and upper-middle-income countries. Across geographical regions, Europe contributed the highest number of publications (95) while the regions of Latin America, Africa, and Australia had the least number of publications; only two each. Of the 113 articles that assessed the rate of occupational disease reporting, 47 studies used data sources from occupational health registries, 44 studies were based on physician reports, 29 studies used data from workers’ compensation claims, and the rest gathered data from a variety of sources including cancer registries and labour surveys.

A total of 89 studies reported on specific occupational diseases, and the most commonly studied conditions were skin diseases (22 studies), cancer (16 studies), respiratory diseases (16 studies), musculoskeletal disorders (11 studies), occupational asthma (9 studies), infectious diseases (7 studies), asbestos-related diseases (5 studies), mental illnesses (4 studies), hearing damage or loss (4 studies), and other chronic conditions (1 study). About 15 studies investigated more than one type of occupational disease. In terms of industry, the health sector had the highest number of studies (10), followed by construction and/or manufacturing (9), agriculture (7), mining and quarrying (3), and other sectors (2). A descriptive summary of the studies included in this review is presented in Appendix 2 in the Supplementary Data section.

### Quality of studies

The majority of the 121 studies (n=116) used a quantitative descriptive research design, followed by qualitative design (n=3), and quantitative non-randomized design (n=2). The quality of the studies was appraised using MMAT with the highest score of 100. Studies that obtained high scores in terms of quality were qualitative studies, with 66.7% (n = 2) scored 100 and 33.3% (n=1) scored 80. Among the quantitative descriptive studies, most (81.9%) had an MMAT score of 80 or higher (n = 94). Only 7 studies (6.0%) scored 40 and 15 studies (12.9%) scored 60. There were only two quantitative non-randomized studies, one scoring 60 and the other scoring 80.

The remaining results are presented according to the study objectives, type of occupational disease, and type of industrial sector. We separated those articles to be analysed for the primary objective, i.e., to estimate the global reporting and underreporting rate of occupational diseases, and the secondary objective of identifying the factors contributing to the underreporting of occupational diseases. Nine articles were assessed for the secondary objective, and the rest were analysed for the primary objective. One article was assessed for both objectives. Of the 113 articles assessed for the primary objective, only two examined underreporting patterns in occupational diseases. Ninety-seven (97) of the 113 studies examined only a specific occupational disease without considering the type of industrial sector. Meanwhile, there were 15 articles which discussed all types of occupational diseases among the general working population. Figure 2 below describes the number of studies in each category.

**Fig 2.**
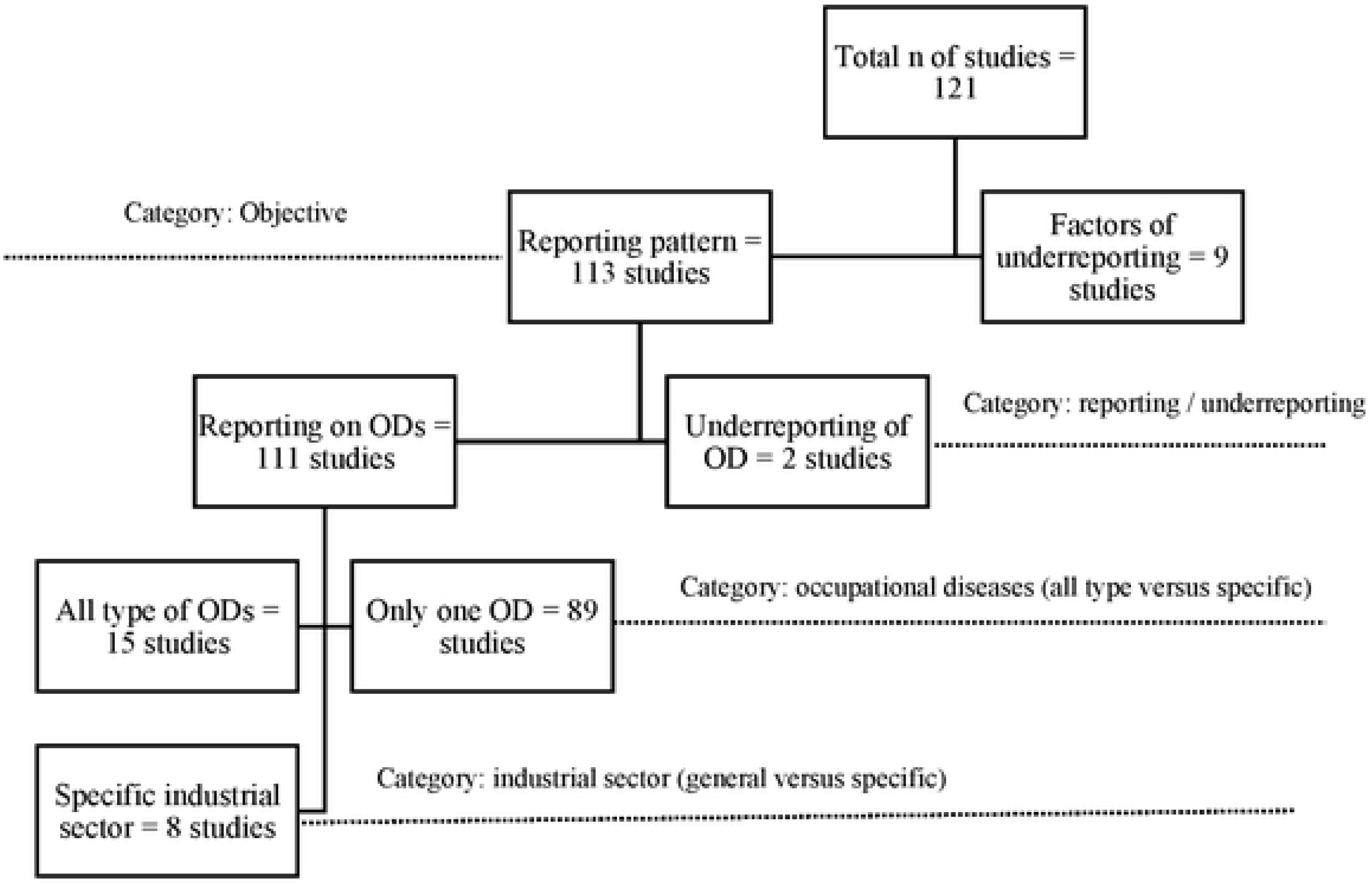
Number of studies in each category based on study objectives, type of occupational diseases and industrial sector.

### Reporting patterns for occupational disease

Data from 15 studies indicate that the number of cases of occupational diseases among the general working population globally ranged from 34 to 37,927 per year with an incidence rate of between 1.71 to 1,387 per 100,000 employees per annum. Almost all the cases were reported in the European region, except for one case that was reported in Asia. The highest rate of occupational disease incidence, as reported by general practitioners between 2006 and 2009, was recorded in the UK. The lowest incidence was recorded in Greece, and it was based on data from workers’ compensation claims. Three specific sectors: agriculture, healthcare, and construction, reported all types of occupational diseases from skin diseases to cancer. The reported annual cases of occupational diseases ranged between 33 to 6,431 in the agricultural sector; 146 to 5,508 in the healthcare sector, and 264 in the construction sector. Table 1 summarises the number of reported cases and incidence rate of occupational diseases in general and specific working populations.

**Table 1.**
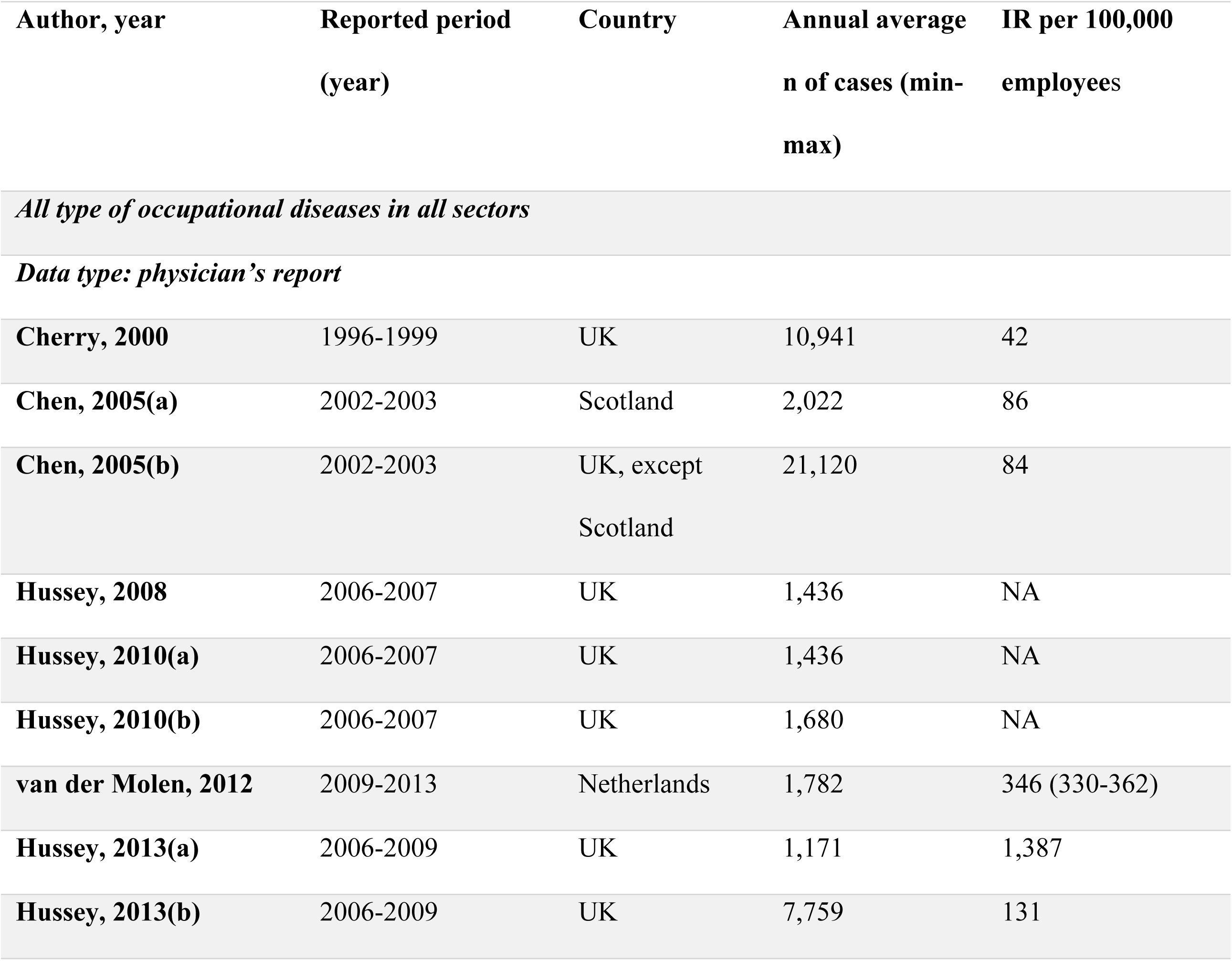
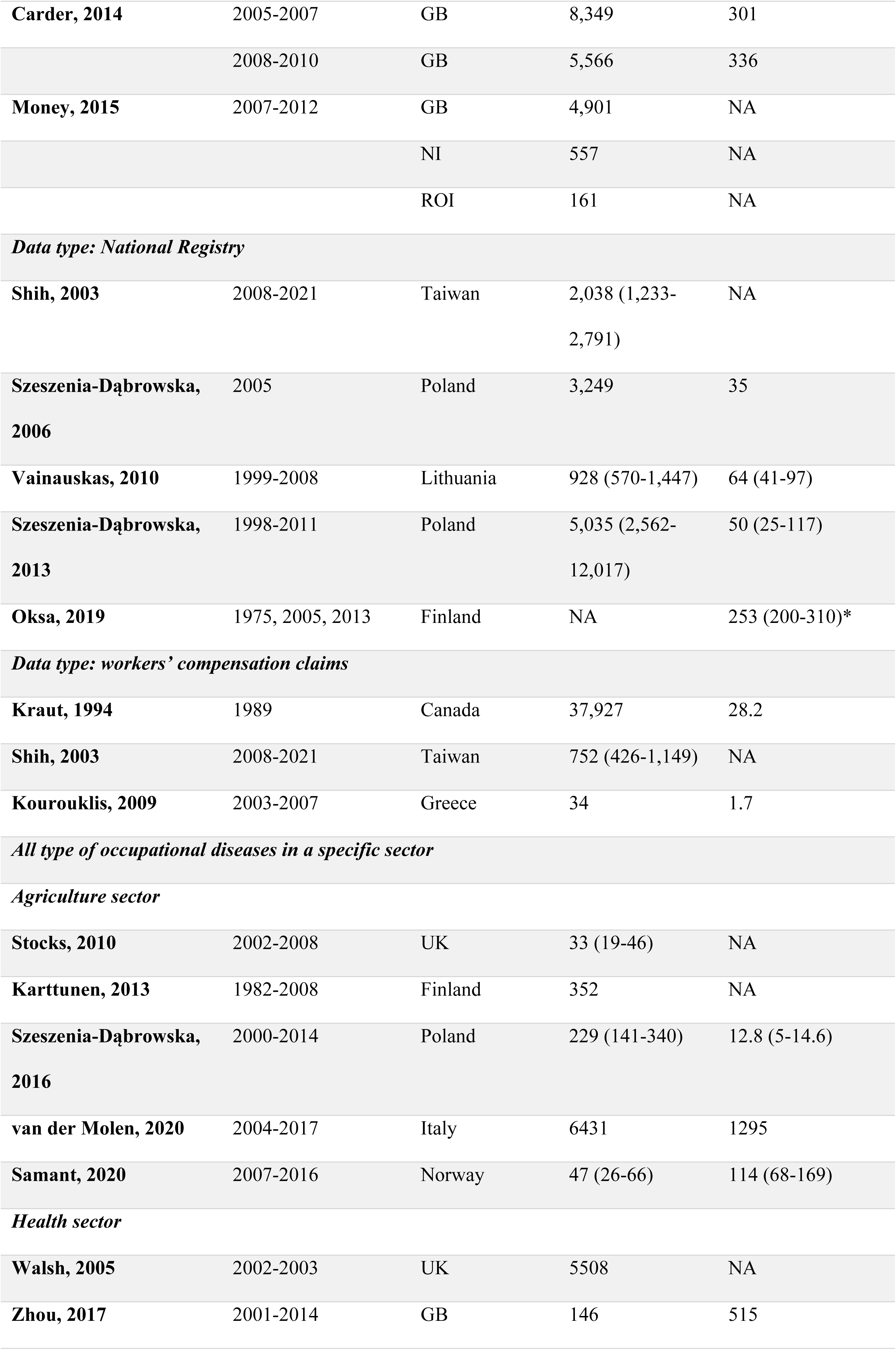
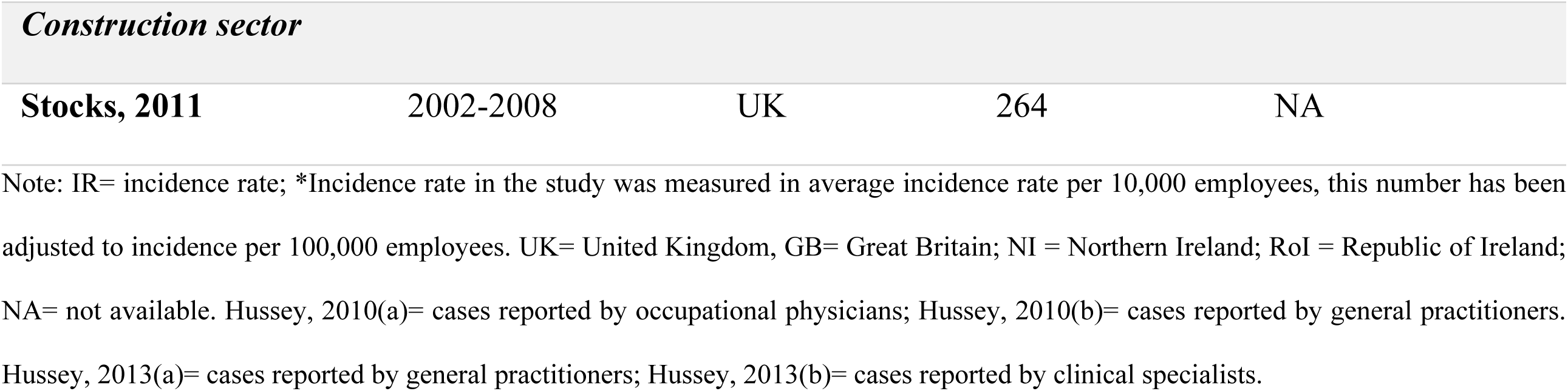
Number of cases and incidence rate of occupational diseases.

### Reporting by type of disease

Occupational-related skin disease was the most reported condition in Europe (18 studies), followed by respiratory diseases (13 studies), cancer (11 studies), musculoskeletal disorders (6 studies), asthma (6 studies), infectious diseases (4 studies), mental illnesses (4 studies), hearing damage/loss (4 studies), and asbestoses (4 studies). Common occupational diseases reported by countries in the European region (UK, Poland, and Finland) included allergic contact dermatitis, irritant contact dermatitis, and contact urticaria as occupational diseases. However, in other regions (Asia, Africa, and Australia), skin diseases were less likely to be studied compared with other types of occupational diseases including musculoskeletal disorders, infectious diseases, and cancer.

Occupational cancer was the second top disease commonly reported in Europe and Asia. The following diagnoses were mostly reported in Europe: skin cancer (4 studies), mesothelioma (3 studies), sinonasal adenocarcinoma (2 studies), and laryngeal cancer (1 study). Many occupational cancers reported in Asia were mesothelioma (South Korean studies), followed by leukemia, and lung cancer (Chinese study).

The average number of reported cases of musculoskeletal disorders was highest in Europe and Asia. Lower back pain was the most frequently reported diagnosis across all musculoskeletal disorders in these two regions. Infectious diseases were also reported in Europe, Asia, and Africa, and were commonly found among healthcare workers. The types of infectious diseases reported among the general working population were diarrheal, scabies, and tuberculosis, while hepatitis B and hepatitis C were frequently found in healthcare workers. Table 2 presents the most reported occupational diseases in the general working population classified by region. We included only studies that reported the number of cases of occupational disease in the general working population.

**Table 2.** Most studied occupational diseases by region.

| Region | Type of occupational disease | n of studies | n of years covered | Average n of annual cases (min-max) |
| --- | --- | --- | --- | --- |
| Europe | Skin diseases | 13 | 6.7 (1-14) | 1,268 (290 – 2,095) |
|  | Respiratory diseases | 6 | 4.6 (1-19) | 1,580 (598 – 3,217) |
|  | Cancer | 11 | 14.2 (3-36) | 160 (4 - 754) |
|  | Musculoskeletal disorders | 5 | 5.6 (2-12) | 1,653 (124 - 4,686) |
|  | Asthma | 5 | 6.6 (3-10) | 335 (97 – 545) |
|  | Infectious diseases | 1 | 4 | 1,402 |
|  | Mental illnesses | 3 | 6.2 (3-14) | 2,788 (526 - 4,709) |
|  | Hearing damage/loss | 2 | 6 (3-9) | 350 (244 – 540) |
|  | Asbestoses | 3 | 23 (20-30) | 58 (6 – 103) |
| Asia | Musculoskeletal disorders | 3 | 13.3 (1-25) | 4,537 (534-9,925) |
|  | Cancer | 3 | 16 (10-23) | 47 (28 - 75) |
|  | Infectious diseases | 1 | 20 | 145 |
|  | Asthma | 2 | 9.7 (6-15) | 30 (14-39) |
|  | Poisonings | 2 | 6.3 (1.6-11) | 645 (60-1,194) |
|  | Asbestoses | 1 | 10 | 7,736 |
|  | Skin diseases | 1 | 1.6 | 68 |
| **Northern America** | Skin diseases | 1 | 1 | 4,289 |
|  | Poisonings | 1 | 11 | 4,932 |
| **Latin America** | Skin diseases | 1 | 6 | 505 |
| **Africa** | Asthma | 1 | 2 | 113 |
|  | Respiratory diseases | 1 | 2 | 1,530 |
|  | Tuberculosis | 1 | 9 | 10 |
| **Australia** | Cancer | 2 | 14 (7-20) | 182 (81-284) |

Note: n of studies defined as number of studies reported that particular occupational disease in the region; n of years covered defined as the average number of years of data collection from all studies; average n of annual cases defined as the average number of cases reported per year, calculated by averaging the total number of cases divided by the number of years of each study. One study might investigate more than one occupational disease and appear more than once in this table. Studies that reported other outcomes, e.g., incident rate, change of incident rate, are not included in the table.

### Reporting of occupational diseases by industrial sector

In our findings, most reports were from health sector, agricultural, construction, mining and quarrying. Studies in the health sector were dominated by infectious diseases (n=7), particularly tuberculosis, hepatitis, and scabies. Three of the seven studies on infectious diseases among healthcare workers were conducted in Germany, one in South Korea, one in South Africa, one in the Czech Republic, and one in Great Britain. One study reported on work-related mental illnesses among healthcare workers. Nurses had a higher number of cases of work-related mental illness compared to doctors. The only study that reported skin disease in healthcare workers was conducted in the Czech Republic. Allergic contact dermatitis, irritant contact dermatitis, and contact urticaria were the most frequently reported cases in the healthcare sector.

In the agricultural sector, we found only two studies: one analysed occupational dermatoses, and the other discussed non-melanoma skin cancers and actinic keratoses. The construction sector had only one study that reported on occupational cancer (i.e. malignant mesothelioma). Musculoskeletal disorders and hearing damage were reported among workers in the mining sector, while skin disease, asthma, and respiratory diseases were reported in the manufacturing sector. Table 3 presents commonly reported occupational diseases across the different industries.

**Table 3.**
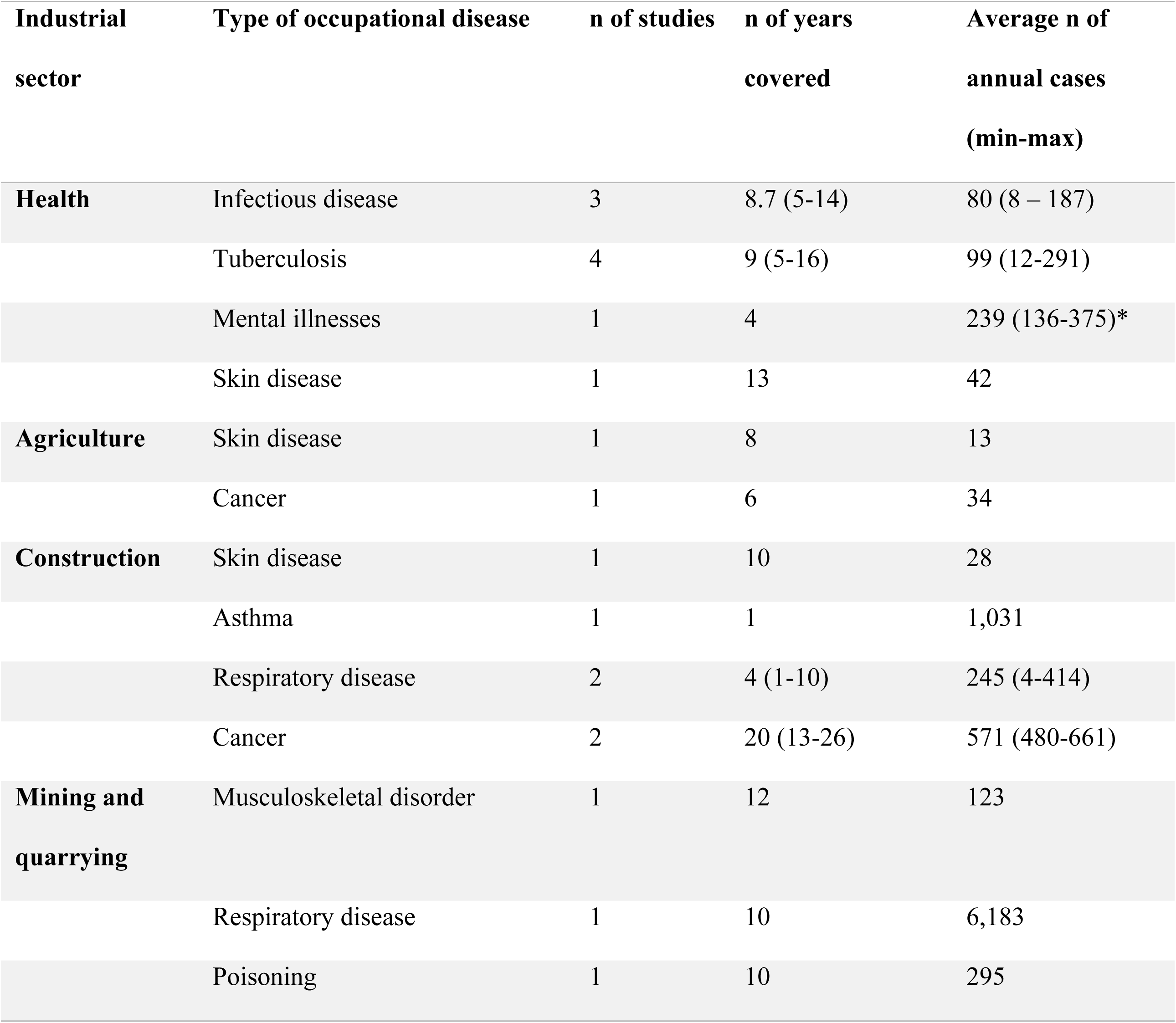
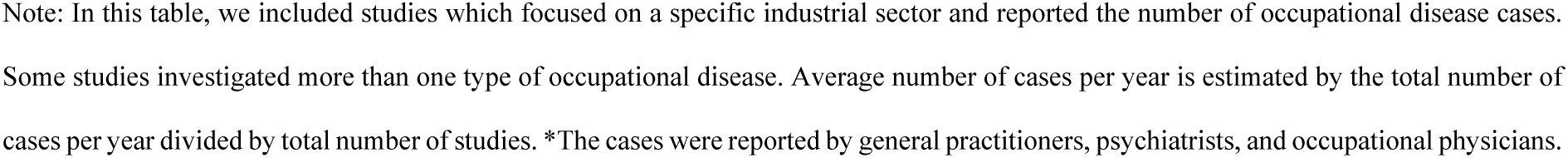
Most reported occupational diseases by industrial sector.

| Industrial sector | Type of occupational disease | n of studies | n of years covered | Average n of annual cases (min-max) |
| --- | --- | --- | --- | --- |
| **Health** | Infectious disease | 3 | 8.7 (5-14) | 80 (8 – 187) |
|  | Tuberculosis | 4 | 9 (5-16) | 99 (12-291) |
|  | Mental illnesses | 1 | 4 | 239 (136-375)* |
|  | Skin disease | 1 | 13 | 42 |
| **Agriculture** | Skin disease | 1 | 8 | 13 |
|  | Cancer | 1 | 6 | 34 |
| **Construction** | Skin disease | 1 | 10 | 28 |
|  | Asthma | 1 | 1 | 1,031 |
|  | Respiratory disease | 2 | 4 (1-10) | 245 (4-414) |
|  | Cancer | 2 | 20 (13-26) | 571 (480-661) |
| **Mining and quarrying** | Musculoskeletal disorder | 1 | 12 | 123 |
|  | Respiratory disease | 1 | 10 | 6,183 |
|  | Poisoning | 1 | 10 | 295 |

Note: In this table, we included studies which focused on a specific industrial sector and reported the number of occupational disease cases. Some studies investigated more than one type of occupational disease. Average number of cases per year is estimated by the total number of cases per year divided by total number of studies. \*The cases were reported by general practitioners, psychiatrists, and occupational physicians.

### Underreporting of occupational diseases

Only two studies investigated the underreporting of occupational diseases at the country level. Both studies estimated a high underreporting rate of between 50% to 95%. One of the two studies (Moreno-Torres 2018) reported the underreporting of all types of occupational diseases, while Skov (1990) estimated the underreporting of occupational-related cancer among the general working population (Table 4).

**Table 4.** Underreporting of occupational diseases.

| Author, year | Reported period (year) | Country | Type of disease | n cases reported | n of cases not reported | Underreporting rate (%) |
| --- | --- | --- | --- | --- | --- | --- |
| Skov, 1990 | 1983-1987 | Denmark | Cancer | 78 | 178 | 50 |
| Moreno-Torres, 2018 | 2000-2015 | Mexico | All | NA | NA | 89 (82-95) |

### Factors affecting the underreporting of occupational diseases

We reviewed seven studies that investigated the underreporting of occupational diseases. Among these studies, one involved in-depth interviews with employees, another was an audit study of employers, and the remaining five were structured surveys targeting of different respondents including physicians, workers, and government representatives. A common theme across all studies was a concern among workers about the possible negative consequences related to job security when reporting occupational diseases. Alaguney et al (24) specifically highlighted this concern among subcontracted workers and those employed without a legal contract. Additionally, limited knowledge of the causal relationship between workplace risks and diseases, as well as lack of awareness of the reporting systems, emerged as key factors influencing the underreporting of occupational diseases (Table 5).

**Table 5.** Factors contributing to the underreporting of occupational diseases.

| Author,<br>year | Country | Type of<br>OD | Method | Type of<br>respondents | Contributing factors |
| --- | --- | --- | --- | --- | --- |
| **Arnaud,**<br>**2010** | France | All | Telephone<br>study | Physicians | • Difficulties in diagnosing ODs<br>• Lack of knowledge of the reporting system<br>• Perceived difficulties in the process of making a claim<br>• Concerns about employee job security |
| **Parhar,**<br>**2011** | Canada | Asthma | Postal<br>survey | Pulmonologists | • Lack of knowledge of the reporting system<br>• Concerns about employee job security |
| **Lysdal,**<br>**2011** | Denmark | Hand<br>eczema | Postal<br>survey | Hairdressers | • Unaware of workplace risks and disease causation<br>• Perceived difficulties in the claiming process<br>• Lack of knowledge of the reporting system |
| **Moldova,**<br>**2017** | 16<br>Eastern | All | Online<br>survey | Official national<br>representatives | • Employer's perceived as loss of revenue |
|  | European countries |  |  |  | • Concerns about employee's job security<br>• Improper monitoring by the authorities |
| **Fagan, 2017** | US | All | Inspections | Employers | • Lack of supervision and training of the onsite medical unit<br>• No clear structure and policies governing the onsite medical unit |
| **Alaguney, 2020** | Turkey | All | Online survey | Physicians | • Employer's perceived as loss of revenue<br>• Concerns about employee's job security, in particular, those working as subcontractor and those without a legal contract |
| **Cheng, 2022** | Taiwan | Asbestos-related diseases | In-depth interviews | Employees | • Unaware of workplace risks and disease causation<br>• Perceived difficulties in the claiming process<br>• Lack of knowledge of the reporting system<br>• Concerns about employee's job security |

## Discussion

This systematic review aimed to estimate the global reporting and underreporting of occupational diseases and to identify factors that influence underreporting. The review included 121 eligible studies, predominantly from high-income countries, with only two articles from upper-middle-income countries, and none from lower-middle-and low-income countries, illustrating the wide gap between high and low/middle-income countries in occupational health research. Europe contributed the highest number of publications in occupational health research; a trend that could be linked to the continent’s high level of industrialization. Historically, the first Industrial Revolution began in the United Kingdom in the 18^th^ century and later spread to other countries (25). This rapid industrialization not only changed the way businesses operate but also raised issues related to occupational health and safety, prompting more industrialized countries to establish laws and regulations to improve working conditions (25).

Despite these advances in high-and upper-middle-income countries, occupational health and safety have remained a lower-priority subject in LMICs compared to other health issues such as infectious diseases, non-communicable diseases, and malnutrition. Our study found the UK to be the most active in contributing to the global evidence on occupational diseases, whereas none of the LMICs included in this review reported the number of occupational disease cases. What was most striking was the limited number of publications from China; despite being the world’s top manufacturing hub (26), we found only two articles on the topic published in Chinese language, with none in English. A previous study identified only six studies on occupational health conducted in LMIC between 1928 and 2019 (27). A key challenge for many LMIC, aside from the gradual rise in industrialization, concerns data quality, completeness, utilization, and limited supporting health information infrastructure (28, 29). Despite having information systems in place, data in many LMICs are frequently reported without accompanying policies or regulations that facilitate data sharing and communication between stakeholders (30). Without a proper system for data collection and interoperability, understanding the extent of occupational disease problems in these countries is difficult.

The working population in LMICs is larger than that of high-income countries (HICs) (31). The United Nations World Population Prospects predicts that by 2030, the largest increases in population will be in LMICs, while populations in HICs will remain relatively stagnant (32). Most of the population in LMICs will be in the productive age group, in contrast to the aging population in HICs. This demographic trend coupled with the growing industrialization in some LIMCs, particularly China, necessitates the establishment of national surveillance systems to prevent and monitor occupational diseases. These systems would require significant investment in funding and human resources (33). They would also require strong political support to make occupational health and safety one of the priority health concerns in LMICs (34). To date, fewer LMICs compared to HICs, have ratified the 2002 Protocol to the Occupational Safety and Health Convention, 1981 (No. 155), which was developed by the International Labour Organization (ILO) (35).

Occupational disease data can be sourced from national registries, physician reports, employers, employees, and workers’ compensation claims. Our findings revealed that many European studies were based on physician reports and/or national registries while in Asia the studies relied more on data from worker compensation claims, and less on physician reports. Reporting of occupational disease is often hindered by a lack of knowledge about workplace risks and their causal relationship with diseases. Many employees have a low awareness of occupational hazards and their health implications (15, 36). Utilizing information collected by physicians could serve as an effective surveillance system, as physicians are better equipped to recognize occupational diseases compared to employers or employees. As our results demonstrate, the UK has an active surveillance system that utilizes reports from general practitioners, occupational physicians, and other medical specialists (rheumatologists, pneumologists, audiologists, and infectious disease specialists). However, not all physicians can easily diagnose occupational diseases due to factors because they had limited training in diagnosing occupational diseases, they might give low priority in recording occupational diseases, or they did not have enough time to identify the occupational hazards in the workplace (37, 38).

Workers face various hazardous risks at their workplace, including exposure to chemical substances, physical hazards, infections, noise, ergonomic stressors, and work stress. In the health sector, we found that biological and psychosocial hazards dominated. This result aligns with the findings of other studies showing that infectious diseases and mental illnesses are the most frequently reported occupational diseases in the health sector (39). Tuberculosis, hepatitis B, and hepatitis C are the most common health concerns among healthcare workers, consistent with previous studies (40–43). In the agricultural sector, workers are exposed to physical, ergonomic, biological, and chemical hazards. They are at risk of developing musculoskeletal disorders, cancer, and dermatitis, as evident from our findings. In terms of the type of diseases according to the region, our study found that non-communicable diseases are the occupational diseases commonly found in America and European countries while in the African region, respiratory and infectious diseases are the most common. These findings align with the regional epidemiology: in America and Europe, where more than 70% of the disease burden is non-communicable, whereas in Africa only 30% is non-communicable (44).

### Strengths and limitations

Data sources were varied and provided little information on the coverage of the working population. There was significant heterogeneity in the methodologies used, particularly in terms of the reporting systems, type of occupational diseases, and specific working population, which restricted our ability to combine data. Consequently, a pooled prevalence of reporting and underreporting of occupational diseases could not be generated using meta-analysis. We included grey literatures, but non-English publications were not included in the analysis. Excluding non-English publications might have little impact on the conclusions(45, 46). Nevertheless, few studies have evaluated the reporting and underreporting trends of occupational diseases using country-level data. To the best of our knowledge, this is the first systematic review summarising the global evidence on the reporting of occupational diseases.

## Conclusions

The evidence indicates a significant gap between high-income countries and LMICs in occupational health research. Occupational diseases are primarily reported in highly developed countries with established national reporting systems. Despite having a larger share of the global working population, LMICs lack evidence on occupational diseases, making it difficult to assess the magnitude of the occupational disease problem. Reporting mechanisms vary substantially with different countries relying on different data sources including worker compensation claims, disease registries, and physician reports. Our findings encourage policymakers, particularly in LMICs, to establish health information infrastructures for occupational disease reporting that enable data sharing and interoperability between stakeholders, including employers, employees, and physicians. Additionally, raising awareness about occupational hazards and improving knowledge about occupational disease reporting are crucial. LMICs would benefit from providing training to employers, employees, and healthcare workers as part of any national awareness campaign.

## Data Availability

All relevant data are within the manuscript and its Supporting Information files.

## Acknowledgements

The authors would like to acknowledge University of New South Wales (UNSW) library for providing methodological guidance and access to scientific journals.

## References

1. WHO/ILO. WHO/ILO joint estimates of the work-related burden of disease and injury, 2000-2016: global monitoring report. Geneva: World Health Organization and International Labour Organization; 2021.

2. WHO. WHO Global Plan of Action on Workers’ Health (2008-2017): Baseline for Implementation. Geneva: World Health Organization; 2013.

3. Difficulties of recording and notification of accidents and diseases in developing countries [press release]. 2019.

4. Organization IL. Challenges for the collection of reliable OSH data 2017 [cited 2022 21 November]. Available from: https://www.ilo.org/wcmsp5/groups/public/---ed_protect/---protrav/---safework/documents/publication/wcms_546702.pdf.

5. Bonnet FV, Joann; Chen, Martha. Women and men in the informal economy: a statistical brief. Manchester; 2019.

6. Boschman JS, Brand T, Frings-Dresen MHW, van der Molen HF. Improving the assessment of occupational diseases by occupational physicians. Occupational Medicine. 2016;67(1):13–9.

7. Verbeek J. When Work is Related to Disease, What Establishes Evidence for a Causal Relation? Saf Health Work. 2012;3(2):110–6.

8. Rushton L. The Global Burden of Occupational Disease. Curr Environ Health Rep. 2017;4(3):340–8.

9. Parhar A, Lemiere C, Beach JR. Barriers to the recognition and reporting of occupational asthma by Canadian pulmonologists. Can Respir J. 2011;18(2):90–6.

10. Spreeuwers D, de Boer AGEM, Verbeek JHAM, van Beurden MM, van Dijk FJH. Diagnosing and reporting of occupational diseases: a quality improvement study. Occupational Medicine. 2008;58(2):115–21.

11. Cegolon L, Lange JH, Mastrangelo G. The Primary Care Practitioner and the diagnosis of occupational diseases. BMC Public Health. 2010;10(1):405.

12. Yamaguchi N, Tazaki H, Okubo T, Toyama T. Periodic urine cytology surveillance of bladder tumor incidence in dyestuff workers. Am J Ind Med. 1982;3(2):139–48.

13. Khoe LC, Mansyur M, Wiseman V, Asante A. What explains the provision of health insurance by Indonesian employers? A trend analysis of the National Labour Force Survey 2018-2022. Health Policy and Planning. 2024;39(7):741–52.

14. Wu Y, Schwebel DC, Hu G. Disparities in Unintentional Occupational Injury Mortality between High-Income Countries and Low- and Middle-Income Countries: 1990⁻2016. Int J Environ Res Public Health. 2018;15(10).

15. Pilusa ML, Mogotlane MS. Worker knowledge of occupational legislation and related health and safety benefits. Curationis. 2018;41(1):e1–e6.

16. Rappin CL, Wuellner SE, Bonauto DK. Employer reasons for failing to report eligible workers’ compensation claims in the BLS survey of occupational injuries and illnesses. American Journal of Industrial Medicine. 2016;59(5):343–56.

17. Green DR, Gerberich SG, Kim H, Ryan AD, McGovern PM, Church TR, et al. Knowledge of work-related injury reporting and perceived barriers among janitors. Journal of Safety Research. 2019;69:1–10.

18. Boadu EF WC, Sunindijo RY. Challenges for occupational health and safety enforcement in the construction industry in Ghana. Construction Economics and Building. 2021;21(1):1–21.

19. Atusingwize E, Musinguzi G, Ndejjo R, Buregyeya E, Kayongo B, Mubeezi R, et al. Occupational safety and health regulations and implementation challenges in Uganda. Arch Environ Occup Health. 2019;74(1-2):58–65.

20. Joshi TK, Bhuva UB, Katoch P. Asbestos ban in India: challenges ahead. Ann N Y Acad Sci. 2006;1076:292–308.

21. Rosenstock L CM, Fingerhut M. Occupational Health. In: Jamison DT BJ, Measham AR, et al, editor. Disease Control Priorities in Developing Countries. New York: The World Bank & Oxford University Press; 2006.

22. Hong QN, Fàbregues S, Bartlett G, Boardman F, Cargo M, Dagenais P, et al. The Mixed Methods Appraisal Tool (MMAT) version 2018 for information professionals and researchers. Education for Information. 2018;34:285–91.

23. 23. Bank TW. World Bank Country and Lending Groups 2021 [Available from: https://datahelpdesk.worldbank.org/knowledgebase/articles/906519-world-bank-country-and-lending-groups.

24. Alaguney ME YA, Demir AU, Ergor OA. Physicians’ opinions about the causes of underreporting of occupational diseases. Archives of Environmental & Occupational Health. 2020;75(3):165–76.

25. Freeman C, Louçã F, Freeman C, Louçã F. 153The British Industrial Revolution: The Age of Cotton, Iron, and Water Power. As Time Goes By: From the Industrial Revolutions to the Information Revolution: Oxford University Press; 2002. p. 0.

26. R B. China is the world’s sole manufacturing superpower: A line sketch of the rise 2024 [updated 17 Jan 2024. Available from: https://cepr.org/voxeu/columns/china-worlds-sole-manufacturing-superpower-line-sketch-rise.

27. Courtice MN OA, Cherrie JW. Less Economically Developed Countries Need Help to Create Healthy Workplaces. Frontiers in Public Health. 2019;7.

28. Hoxha K, Hung YW, Irwin BR, Grépin KA. Understanding the challenges associated with the use of data from routine health information systems in low-and middle-income countries: A systematic review. Health Information Management Journal. 2022;51(3):135–48.

29. Abdul-Rahman T, Ghosh S, Lukman L, Bamigbade GB, Oladipo OV, Amarachi OR, et al. Inaccessibility and low maintenance of medical data archive in low-middle income countries: Mystery behind public health statistics and measures. Journal of Infection and Public Health. 2023;16(10):1556–61.

30. 30. O‘Neil S, Taylor S, Sivasankaran A. Data Equity to Advance Health and Health Equity in Low- and Middle-Income Countries: A Scoping Review. DIGITAL HEALTH. 2021;7:20552076211061922.

31. King EM, Randolph HL, Floro MS, Suh J. Demographic, health, and economic transitions and the future care burden. World Dev. 2021;140:105371.

32. United Nations Department of Economic and Social Affairs PD. World Population Prospects 2022: Summary of Results United Nations; 2022.

33. Witter S, Hamza MM, Alazemi N, Alluhidan M, Alghaith T, Herbst CH. Human resources for health interventions in high- and middle-income countries: findings of an evidence review. Human Resources for Health. 2020;18(1):43.

34. Mills A. Health Care Systems in Low- and Middle-Income Countries. New England Journal of Medicine. 2014;370(6):552–7.

35. Protocol of 2002 to the Occupational Safety and Health Convention, 1981, (2002).

36. Geleta DH, Alemayehu M, Asrade G, Mekonnen TH. Low levels of knowledge and practice of occupational hazards among flower farm workers in southwest Shewa zone, Ethiopia: a cross-sectional analysis. BMC Public Health. 2021;21(1):232.

37. Gök AS, Yilmaz TE, Kasim İ, Şencan İ, Özkara A. Occupational Health and Disease Knowledge and Practices of Family Physicians. J Occup Environ Med. 2020;62(11):e625–e9.

38. Holness DL, Kudla I, Brown J, Miller S. Awareness of occupational skin disease in the service sector. Occupational Medicine. 2016;67(4):256–9.

39. Vecchio D, Sasco AJ, Cann CI. Occupational risk in health care and research. Am J Ind Med. 2003;43(4):369–97.

40. Mohanty A, Kabi A, Mohanty AP. Health problems in healthcare workers: A review. J Family Med Prim Care. 2019;8(8):2568–72.

41. O’Hara LM, Yassi A, Zungu M, Malotle M, Bryce EA, Barker SJ, et al. The neglected burden of tuberculosis disease among health workers: a decade-long cohort study in South Africa. BMC Infectious Diseases. 2017;17(1):547.

42. Kacem M, Dhouib W, Bennasrallah C, Zemni I, Abroug H, Ben Fredj M, et al. Occupational exposure to hepatitis C virus infection and associated factors among healthcare workers in Fattouma Bourguiba University Hospital, Tunisia. PLoS One. 2022;17(9):e0274609.

43. Westermann C, Peters C, Lisiak B, Lamberti M, Nienhaus A. The prevalence of hepatitis C among healthcare workers: a systematic review and meta-analysis. Occupational and Environmental Medicine. 2015;72(12):880.

44. Vos T, Lim SS, Abbafati C, Abbas KM, Abbasi M, Abbasifard M, et al. Global burden of 369 diseases and injuries in 204 countries and territories, 1990&#x2013;2019: a systematic analysis for the Global Burden of Disease Study 2019. The Lancet. 2020;396(10258):1204–22.

45. Dobrescu AI, Nussbaumer-Streit B, Klerings I, Wagner G, Persad E, Sommer I, et al. Restricting evidence syntheses of interventions to English-language publications is a viable methodological shortcut for most medical topics: a systematic review. Journal of Clinical Epidemiology. 2021;137:209–17.

46. Nussbaumer-Streit B, Klerings I, Dobrescu AI, Persad E, Stevens A, Garritty C, et al. Excluding non-English publications from evidence-syntheses did not change conclusions: a meta-epidemiological study. Journal of Clinical Epidemiology. 2020;118:42–54.

